# Moderating the link between discrimination and adverse mental health outcomes: Examining the protective effects of cognitive flexibility and emotion regulation

**DOI:** 10.1101/2023.02.11.23285777

**Authors:** Yutong Zhu, Wisteria Deng

## Abstract

Discrimination is associated with mental health problems. While prior research has demonstrated the significance of emotion regulation in explaining the onset and development of discrimination-related anxiety, few studies investigated this dynamic with cognitive flexibility among sexual and/or racial minority individuals. The current study incorporated cognitive flexibility to investigate its potential buffering effects on discrimination-related anxiety. 221 individuals, 37.6% of whom (*n* = 83) identified as sexual and/or racial minorities, responded to an online questionnaire about their levels of cognitive flexibility and emotion regulation, perceived discrimination, and anxiety. Moderated mediation analyses were conducted with these variables. Our findings indicated that emotion regulation difficulty (ERD) mediated the relationship between discrimination and anxiety, while cognitive flexibility had a strong moderating effect on the relationship between ERD and anxiety. These results suggested new research directions and implied the therapeutic potential of advancing cognitive flexibility skills with emotion regulation training in anxiety prevention and treatments. Future research is needed to investigate cognitive flexibility as a transdiagnostic mechanism underlying the onset and development of anxiety, to potentially lead to novel prevention or intervention for marginalized people facing additional stressors like discrimination.

## Introduction

Discrimination incurs substantial costs as it associates with negative health outcomes [1]. Discrimination can lead to stress-related difficult emotions such as sadness [2], anger [3], and grief [4]. These negative emotions can result in adverse physical and behavioral responses, including cardiovascular diseases [5], sleep difficulties [6], and substance abuse [7]. It is especially harmful to racial and/or sexual minorities, who may experience low self-esteem, anxiety, and stress-related disorders as a result of discrimination [8-11]. Asian Americans reported a marked increase in racial discrimination during the COVID-19 pandemic, which led to worsened anxiety and depression symptoms [12]. Additionally, a nationally representative survey found that 36% of LGBTQ adults reported experiencing discrimination, with 52% reporting negative impacts on their mental well-being [13]. Considering the discrimination experienced by minority populations and subsequent adverse mental health outcomes, it is important to identify potential risk and protective factors to inform public health decisions and intervention designs for minority individuals.

Given that the association between discrimination and adverse mental health outcomes is fueled by emotional responses, emotion regulation may moderate this relationship. Previous literature identifies emotion regulation difficulty (ERD) as a risk factor underlying the development of stress-related disorders. Emotion regulation refers to one’s explicit or implicit control over emotional states [14]. ERD may manifest as avoidance or suppression of emotional experiences and can lead to persistent worry and an excessive focus on emotional cues, which contribute to the development of generalized anxiety disorder [15-16]. Another study has found that ERD is directly linked to anxiety diagnoses even after controlling for worry, trait anxiety, and depressive symptoms [17].

Apart from affective factors, cognitive mechanisms may also influence anxiety levels after experiencing discrimination. While previous research has demonstrated that ERD can lead to increased anxiety, little has been explored regarding how cognitive flexibility can interact with emotion regulation to alleviate adverse mental health outcomes. Cognitive flexibility refers to the ability to adjust one’s beliefs and/or behaviors to better adapt to the environment [18]. People with a higher level of cognitive flexibility can disengage from the previous mindsets to incorporate new information and form new beliefs that are more appropriate to the current context. High cognitive flexibility reflects high cognitive control and the ability to reevaluate current situations (e.g., cognitive reappraisal); these cognitive abilities and skills are shared by adaptive emotion regulation strategies [19-20]. Thus, it can be hypothesized that cognitive flexibility may interact with emotion regulation ability to buffer against anxiety incurred by discrimination. Meanwhile, employing maladaptive emotion regulation techniques can result in low moods and reduced motivation to revise existing beliefs, consequently negatively impacting cognitive flexibility [21]. A better understanding of the interplay between emotion regulation and cognitive flexibility is crucial to understand the development of negative mental health impacts of discrimination.

### Current study

The current study aimed to examine the interaction between cognitive flexibility and emotion regulation difficulties (ERD) in relation to discrimination-induced anxiety. We hypothesized that ERD would mediate the relationship between discrimination and anxiety. Additionally, cognitive flexibility would moderate the association between discrimination and anxiety as well as the link between ERD and anxiety (i.e., these associations will be weaker for individuals with high cognitive flexibility) (S1 Fig). By exploring the interplay between emotion regulation and cognitive flexibility, this study aims to uncover potential protective factors against discrimination-related anxiety. Our findings may provide valuable insights for public health recommendations and innovative interventions for minority groups who experience prevalent discrimination and heightened anxiety.

## Materials and methods

### Procedure and participants

Participants (*N* = 221, *M*_*age*_ = 41.18, *SD*_*age*_ = 12.09) were recruited via Amazon’s Mechanical Turk (MTurk), an online crowdsourcing platform that provides access to a large and diverse sample for mental health research studies (demographics see Table 1). MTurk users who live in the United States and are 18 years or older were eligible to participate in this study. According to guidelines for research using crowdsourced samples, the study exclusively recruited participants who have a history of submitting high-quality answers [22]. All participants need to complete at least 500 MTurk studies, with 98% of those studies accepting their responses. Participants received $6 for study compensation.

**Table 1.** Demographic Characteristics of Participants.

| Gender | # | % | Psychiatric History | # | % |
| --- | --- | --- | --- | --- | --- |
| Male<br>($M_{age} = 41.25$, $SD = 12.56$) | 115 | 52.04% | Yes | 55 | 24.89% |
| Female<br>($M_{age} = 41.09$, $SD = 11.61$) | 106 | 47.96% | No | 166 | 75.11% |
| Sexual Orientation |  |  | Psychiatric Medication History |  |  |
| Heterosexual | 182 | 82.35% | Yes | 47 | 21.27% |
| Homosexual | 5 | 2.26% | No | 174 | 78.73% |
| Bisexual/Pansexual | 26 | 11.76% | Current Medication (if applicable) |  |  |
| Asexual/Aromantic | 7 | 3.17% | Mental health | 33 | 56.90% |
| Queer/Questioning | 1 | 0.45% | Medical problems | 27 | 46.55% |
| Race/Ethnicity |  |  | General health | 3 | 5.17% |
| White/Caucasian | 167 | 75.57% | Family History of Mental Illness |  |  |
| Black/African American | 23 | 10.41% | Yes | 61 | 27.60% |
| Asian | 15 | 6.79% | No | 160 | 72.40% |
| Native American/Indigenous | 1 | 0.45% | Employment |  |  |
| Others | 15 | 6.79% | Currently employed, full-time | 159 | 71.95% |
| Hispanic/Latino | 32 | 14.48% | Currently employed, part-time | 28 | 12.67% |
| Marital Status |  |  | Student | 3 | 1.36% |
| Single/Never married | 78 | 35.29% | Retired | 7 | 3.17% |
| Currently married | 88 | 39.82% | Unable to work | 3 | 1.36% |
| Divorced | 21 | 9.50% | Stay at home parent | 5 | 2.26% |
| Separated | 2 | 0.90% | Currently unemployed, looking for employment | 11 | 4.98% |
| Currently in a relationship but not married | 32 | 14.48% | Currently unemployed, not looking for unemployment | 5 | 2.26% |
| Educational Level |  |  | Household Income |  |  |
| Less than high school | 1 | 0.45% | Less than \$20,000 | 21 | 9.50% |
| High school | 15 | 6.79% | \$20,000 to \$34,999 | 20 | 9.05% |
| Some college | 40 | 18.10% | \$35,000 to \$49,999 | 45 | 20.36% |
| 2-year degree/Associate's degree | 28 | 12.67% | \$50,000 to \$74,999 | 53 | 23.98% |
| 4-year degree/Bachelor's degree | 105 | 47.51% | \$75,000 to \$99,999 | 38 | 17.19% |
| Master's degree | 27 | 12.22% | \$100,000 to \$149,999 | 28 | 12.67% |
| Doctorate | 5 | 2.26% | \$150,000 to \$199,999 | 9 | 4.07% |
| Native Language | | | \$200,000 or more | 7 | 3.17% |
| English | 218 | 98.64% |  |  |  |
| Others | 3 | 1.36% |  |  |  |

### Measures

#### Minority status

Participants were asked to report their race and sexual orientation. We computed a binary variable called “minority status”: participants were coded 1 (“yes”) if they reported being either non-heterosexual or non-White. Participants reported to be both heterosexual and White were coded 0 (“no”) in this variable.

#### Discrimination

Everyday Discrimination Scale (EDS) is a 9-item scale that measures the frequency of day-to-day microaggression or discrimination [23]. Each item contains a four-point Likert scale ranging from 0 (“never”) to 5 (“almost every day”). The EDS also contains a follow-up question to ask the respondent about their perceived “main reasons” for their discriminative experiences (e.g., race, gender, sexual orientation, etc.). The EDS has been shown to have high construct validity among racial minority groups [24].

#### Cognitive flexibility

The Interpretive Inflexibility Task (IIT) is a picture-based scenario task developed from the Emotional BADE task [25-26]. In contrast to the verbal scenarios used in the Emotional BADE task, each of the 24 IIT scenarios is based on a photograph of an interpersonal situation and is progressively presented to the respondents, with 12 leading to a positive resolution and 12 leading to a negative resolution. Each IIT scenario is presented to the respondent three times: with 80%, 20%, and 0% of the photograph blurred at each time. The blurred areas were chosen to conceal the emotional valence of the scenario. By gradually reducing the percentage of blurred photos, respondents are provided with more information, which may help to resolve the initial blurring. The IIT produces an interpretation bias index reflecting moment-to-moment fluctuations in interpretation bias (1), with a high interpretation bias index indicating high flexibility in revising initial biased interpretations. Scenarios that lead to a positive resolution were scored for positive flexibility, whereas scenarios that lead to a negative resolution were scored for negative flexibility [25].

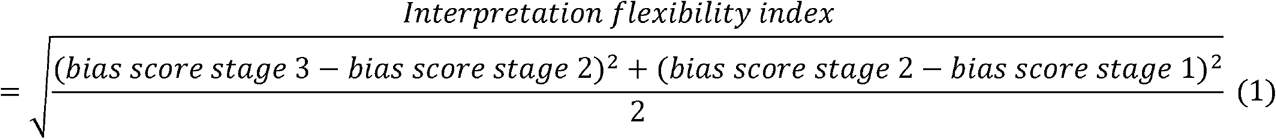

#### Emotion regulation difficulty (ERD)

The Difficulties in Emotion Regulation Scale (DERS) is a self-report assessment designed to measure an individual’s trait-level emotion regulation capabilities. The DERS consists of 36 items, with higher scores indicating greater difficulties in emotion regulation. This scale assesses an individual’s perceived ability to regulate their emotions, including their awareness of emotions, acceptance of emotions, and ability to adjust cognitively and behaviorally during times of emotional distress [27].

#### State anxiety

State-Anxiety Inventory (STAI) is a commonly used measure of trait and state anxiety [28]. STAI has been widely used in clinical settings for anxiety disorder diagnoses, as it can sensitively distinguish anxiety from depressive symptoms. We used the short version of the measurement (STAI-5) developed by Zsido and his collogues [29]. STAI-5 contains 5 items for trait anxiety and 5 items for state anxiety. The short version of STAI has been found to have comparable psychometric properties compared to the 40-item long version [29]. Higher scores indicate a greater level of anxiety.

### Statistical analysis

Statistical analyses were performed using SPSS Mac Version 26 (SPSS Science, Chicago, IL, United States). Descriptive analyses were first performed to investigate any potential correlations among the main study variables. Independent t-tests were conducted comparing the level of discrimination concerning socio-demographic status. SPSS Macro PROCESS Version 4.1 was used for the moderated mediation analysis [30]. Analytical procedures of Model 15 instead of 14 were adopted to investigate the potential moderating effects on the direct and indirect effects. As in the hypothesized model, daily discrimination was the independent variable and state anxiety was the dependent variable. ERD was included as the mediator of the relationship between daily discrimination and anxiety. Positive and negative flexibility were separately analyzed as the moderator of the relationship between daily discrimination and ERD, and anxiety respectively. In addition, an alternative model with cognitive flexibility as the mediator and ERD as the moderator would also be tested to investigate the potential reciprocal influence of emotion regulation and cognitive flexibility. All variables were centered on the mean to avoid multicollinearity [31]. We used 5,000 bootstrap samples and determined the mediation and moderation effects of the 95% confidence interval. Moreover, to better account for the moderating effects [32], we examined the conditional indirect effects of the moderators at -1SD, the mean, and +1SD. Statistical significance was set at a two-tailed p-value < .05.

## Results

### Descriptive statistics

Table 2 presents the descriptive statistics and correlations for the main study variables. Reported discrimination was strongly correlated with all other variables: minority status (*r* = .190, *p* = .005), positive flexibility (*r* = -.231, *p* = .001), negative flexibility (*r* = -.189, *p* = .005), state anxiety (*r* = .389, *p* < .001), and ERD (*r* = .314, *p* < .001). ERD was strongly correlated with state anxiety (*r* = .498, *p* < .001). Positive and negative flexibility strongly correlated with each other (*r* = .544, *p* < .001).

**Table 2.** Descriptive statistics and correlations of study variables.

| Variables | *M* | *SD* | 1 | 2 | 3 | 4 | 5 | 6 |
| --- | --- | --- | --- | --- | --- | --- | --- | --- |
| 1. Minority Status | .38 | .485 | - |  |  |  |  |  |
| 2. Positive Flexibility | .728 | .438 | -.138* | - |  |  |  |  |
| 3. Negative Flexibility | 1.034 | .569 | -.154* | .544*** | - |  |  |  |
| 4. State Anxiety | 6.28 | 2.373 | .129 | -.344*** | -.284*** | - |  |  |
| 5. ERD | 77.82 | 25.464 | .130 | -.154* | -.072 | .498*** | - |  |
| 6. Discrimination | 20.51 | 11.112 | .190** | -.231** | -.189** | .389*** | .314*** | - |

$N = 221$ , \* $p < 0.05$ , \*\* $p < 0.01$ , \*\*\* $p < 0.001$ . ERD: Emotion Regulation Difficulty

### Socio-demographic data and discrimination

There were significant differences in discrimination with respect to minority status. Consistent with previous literature, racial and/or sexual orientational minority individuals are more likely to experience discrimination (*t* = -2.870, *p* = .005). Mean comparison showed no significant associations between discrimination and gender (*t* = -.415, *p* = .679), psychiatric history (*t* = .715, *p* = .475), marital status (*t* = 1.118, *p* = .349), educational level (*t* = .350, *p* = .910), and household income (*t* = 1.894, *p* = .080).

### Cognitive flexibility and emotion regulation’s moderated mediating effects on state anxiety

When including flexibility in endorsing positive outcomes (i.e., positive flexibility) as the moderator, the results suggested a significant impact of discrimination on ERD (*b* = .720, *t* = 4.899, *p* < .001), and ERD had a significant impact on state anxiety (*b* = .074, *t* = 7.624, *p* < .001). Accounting for the mediating effect of ERD, discrimination did not have a significant impact on state anxiety (*b* = .035, *t* = 1.517, *p* = .131). Additionally, positive flexibility significantly moderated the indirect effect of ERD on anxiety (*b* = -.054, *t* = -4.753, *p* < .001, ΔR^2^ = .060), but did not moderate the direct effect of discrimination on anxiety (*b* = .000, *t* = -.008, *p* = .993). Bootstrap testing also indicated a moderating effect on indirect effect. Specifically, individuals with low positive flexibility observed a stronger relationship between ERD and state anxiety, whereas those with high positive flexibility are less likely to develop anxiety following ERD. In sum, emotion regulation influenced the relationship between discrimination and anxiety, while positive flexibility moderated the relationship between emotion regulation and anxiety (Fig 1).

**Fig 1.**
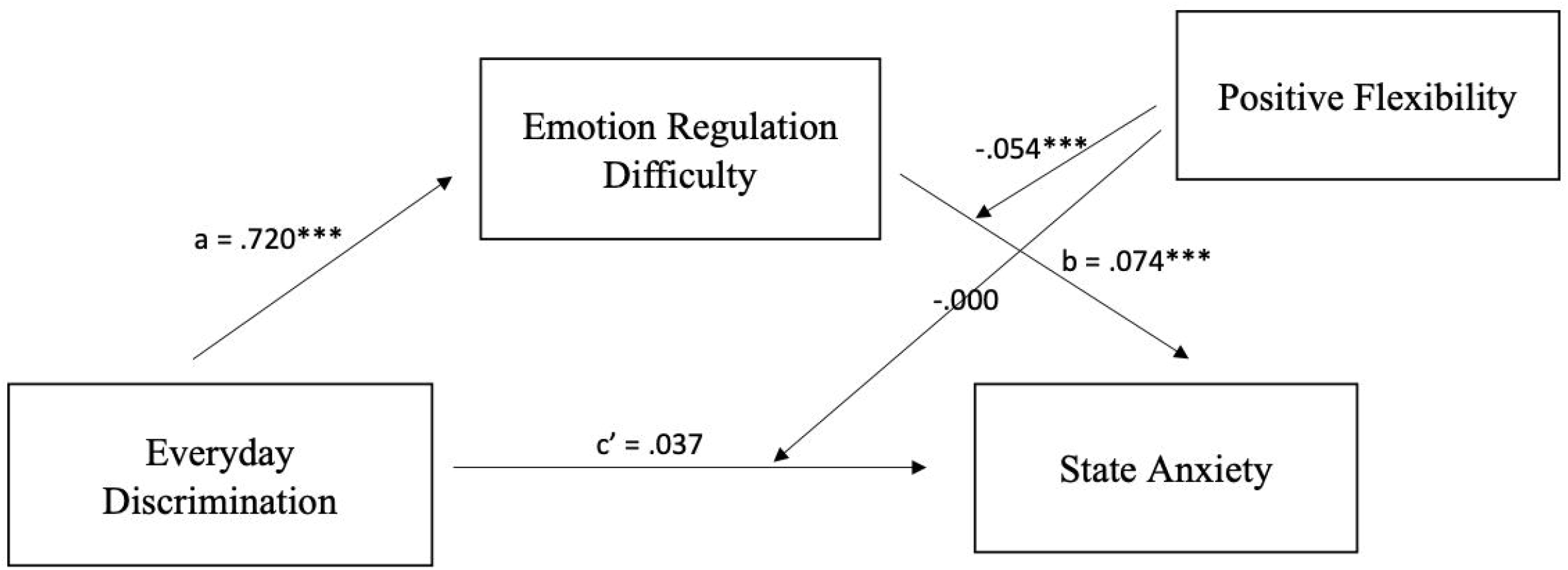
The standardized regression coefficients for the mediating effect of ERD and the moderating effect of positive flexibility on the relationship between discrimination and anxiety. \**p* < .05, \*\**p* < .01, \*\*\**p* < .001.

Similarly, flexibility in endorsing negative outcomes (i.e., negative flexibility) also moderated the association between ERD and state anxiety. Results indicated that discrimination had a significant impact on ERD (*b* = .720, *t* = 4.899, *p* < .001) and ERD had a significant impact on state anxiety (*b* = .074, *t* = 6.010, *p* < .001). Unlike when positive flexibility is the moderator, discrimination still had a significant impact on state anxiety after being mediated by ERD (*b* = .063, *t* = 2.207, *p* = .028). Negative flexibility significantly moderated the impact of ERD on anxiety (*b* = -.034, *t* = -3.392, *p* = .001) but did not moderate the impact of discrimination on anxiety (*b* = -.024, *t* = -.965, *p* = .336). Bootstrap testing indicated a moderating effect on indirect effect and the change of effect size (ΔR^2^) was .031. In other words, individuals with high ERD showed a stronger relationship between discrimination and state anxiety, while high negative flexibility weakened the relationship between ERD and state anxiety (Fig 2).

**Fig 2.**
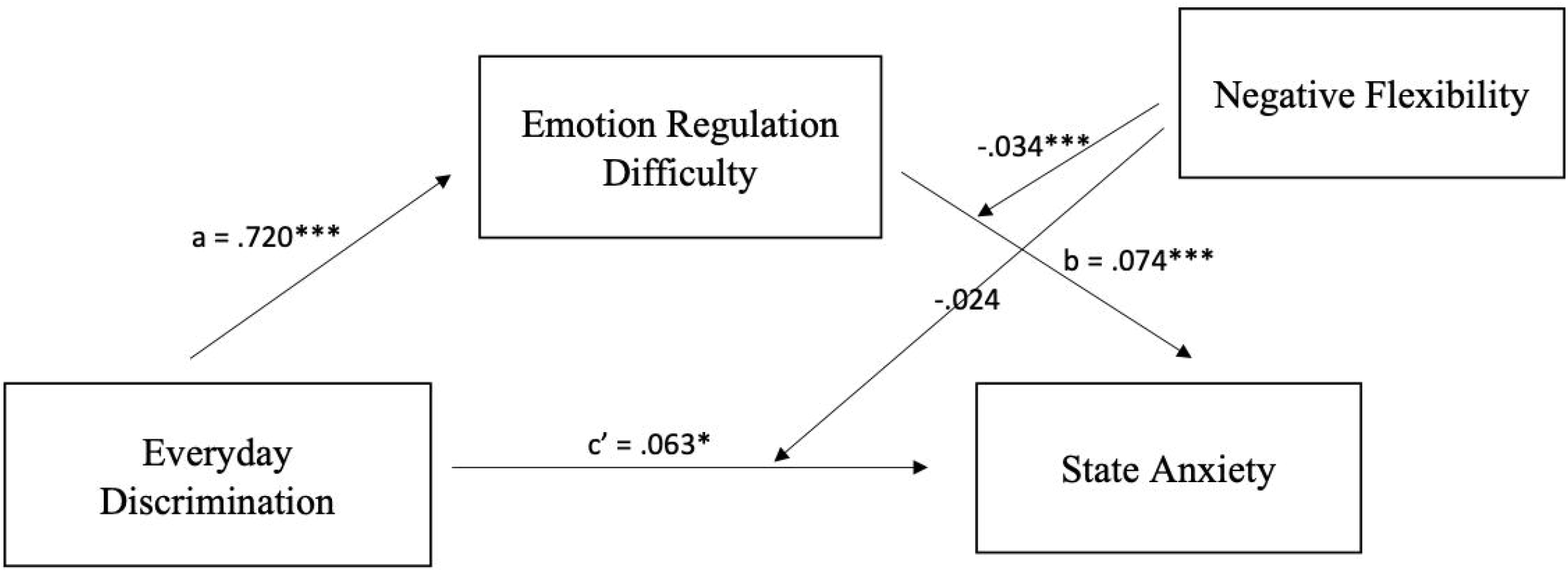
The standardized regression coefficients for the mediating effect of ERD and the moderating effect of negative flexibility on the relationship between discrimination and anxiety. \**p* < .05, \*\**p* < .01, \*\*\**p* < .001.

Moreover, because of the limitation of cross-sectional data, causation and directionality cannot be inferred among the variables. Thus, we tested alternative models using ERD as the moderator and cognitive flexibility as the mediator. The results indicated that positive and negative flexibility partially mediated the relationship between discrimination and state anxiety. ERD moderated both the direct and indirect effects when negative flexibility was the moderator, but only moderated the indirect effect when positive flexibility was the moderator (S2 and S3 Figs for coefficients). Thus, cognitive flexibility and emotion regulation reciprocally influence each other while shaping the relationship between discrimination and state anxiety.

## Discussion

The current study investigated the effects of emotion regulation and cognitive flexibility on discrimination-related anxiety. Our findings confirmed that the reported discrimination level is higher among sexual and racial minorities. ERD mediated the relationship between discrimination and anxiety, while cognitive flexibility (in endorsing both positive and negative outcomes) moderated the positive association between ERD and anxiety.

Consistent with the previous literature [33-35], our findings highlighted the significance of ERD in explaining discrimination-related anxiety, suggesting the importance of implementing adaptive emotion regulation strategies for sexual and/or racial minority individuals. Past neuroimaging results revealed the neural correlates supporting the association between ERD and anxiety. fMRI studies indicated that ERD arises from dysregulation in the amygdala, as well as abnormalities in its connectivity with the frontal-cortical areas [36]. Elevated amygdala activation was found in at-risk individuals without an anxiety disorder diagnosis [37]. Moreover, negative amygdala-vmPFC connectivity was associated with social anxiety disorder symptomatology, and effective treatments that improve ERD often reduce this brain abnormality [38]. Another study found that emotion dysregulation relates to a reduction in amygdala-rVLPFC functional connectivity among female adolescents, and this hypoconnectivity predicted anxiety symptoms during nine-month follow-ups [39]. Overall, supported by neuroimaging findings, emotion regulation difficulty is a risk factor for the onset, development, and maintenance of anxiety.

Also, we pointed out the importance of cognitive flexibility in buffering against the association between ERD and anxiety and subsequently between discrimination and anxiety. These findings emphasized the importance of incorporating cognitive flexibility training in disseminating emotion regulation strategies. While existing anxiety interventions targeting emotion dysregulation often train individuals on transferrable skills that can also improve cognitive flexibility, few of them structurally include cognitive flexibility as an intervention target. For example, anxiety prevention programs for children and adolescents included cognitive-behavioral training such as cognitive restructuring (i.e., the ability to notice negative thinking patterns) [40]. Mennin and Fresco’s Emotion Regulation Therapy (ERT) contains behavioral intervention on regulatory skills such as attentional flexibility and cognitive reframing skills (i.e., belief flexibility) [41]. However, none of these intervention frameworks explicitly draw the link between cognitive flexibility and emotion regulation ability. Given our findings on how cognitive flexibility may interact with emotion regulation to protect against anxiety, it may be helpful to clearly include cognitive flexibility as a goal of the intervention. Ultimately, emotion regulation presents a form of flexibility in modifying one’s emotions, which falls in line with the cognitive flexibility required to revise existing beliefs and thoughts. Drawing connections between these different forms of flexibility may bring about insight, increase the effectiveness of anxiety interventions, and improve treatment outcomes.

Also, it is important to note that the direct effect of discrimination on anxiety was significant when negative flexibility (*p* =.028), but not positive flexibility (*p* =.131), was the moderator. In parallel, moderation analyses indicated that the impact of ERD on anxiety is greater when anxiety is explained by positive flexibility (ΔR^2^ = .060) than negative flexibility (ΔR^2^ = .031). These findings supported prior work showing that negative and positive flexibility have differential effects in predicting affective symptoms [25-26], further confirming the distinct effects of negative and positive flexibility in explaining anxiety. Compared to negative flexibility, positive flexibility may be more closely related to the association between emotion regulation and anxiety. Anxiety disorders are characterized by negative interpretation bias, or the tendency to interpret ambiguous situations as a negative or catastrophic solution [42]. Therefore, the ability to flexibly revise biased interpretations in a positive direction may rely more on effective emotion regulation skills and has salient impacts on anxiety outcomes. To further evaluate the mechanisms of positive and negative flexibility, future research needs to examine the affective component of biased interpretations and the corresponding affective processing during belief revision in greater detail.

Furthermore, our alternative model using cognitive flexibility as the mediator and ERD as the moderator suggested that cognitive flexibility partially mediated the relationship between discrimination on anxiety, while ERD moderated this mediation. Altogether, our findings showed that the interplay of cognitive and affective revisioning processes goes in both directions. Indeed, emotion regulation and cognitive flexibility interact with each other dynamically during information processing and together influences the development of discrimination-induced anxiety, as each moderate the impact of the other. This finding reiterated the therapeutic implications of including cognitive flexibility training in anxiety prevention and treatments to improve outcomes. Moreover, considering the transferrable skills between emotion regulation and cognitive flexibility (e.g., cognitive reappraisal), cognitive flexibility can also be used as an indicator to evaluate the effectiveness of anxiety interventions that target emotion dysregulation.

The study is not without limitations. First, although the study had a clear objective to include sexual and racial minorities in the sampling procedure, the online design still restricted our ability to gather a representative sample. Future research should incorporate in-person participant recruitment to make sure that minority populations with diverse identities are included. Second, the cross-sectional nature of this study does not allow for the inference of causal relationships between variables. In fact, our results indicated that cognitive flexibility and ERD were interchangeable as mediators and moderators when explaining the association between discrimination and anxiety, which further suggests the reciprocal influences of these factors during affective processing. Thus, future research can use real-time data collection methods, such as ecological momentary assessments (EMAs), to capture the temporal variations paralleling individuals’ cognitive and affective processing [43]. By investigating the dynamics of cognitive flexibility and emotion regulation under real-world affective contexts, scholars would have a more comprehensive understanding of how affective and cognitive factors interact in predicting anxiety.

## Conclusion

Previous research suggested the significance of emotion regulation in explaining the onset and development of discrimination-related anxiety. The current study further included cognitive flexibility in this dynamic. We focused on individuals’ ability to update existing beliefs and found a significant moderated mediating impact of cognitive flexibility and ERD on the association between discrimination and anxiety. ERD mediated the connection between discrimination and anxiety, with higher difficulties indicating a higher level of anxiety, while both positive and negative flexibility buffered against the negative impact of ERD. Alternative model analyses suggested the possible reciprocal influence of cognitive flexibility and emotion regulation in influencing discrimination-related anxiety. Despite the limitation of the sample diversity and cross-sectional study design, these findings foreshadowed novel research directions and implied therapeutic applications. Future studies like this will advance our understanding of anxiety mechanisms and potentially lead to novel prevention and interventions for marginalized individuals facing additional stressors like discrimination.

## Data Availability

Data is only available on request.

## Acknowledgments

The authors declare no conflict of interest and the authors received no funding from an external source.

## Supporting information

**S1 Fig. Hypothesized Model**.

**S2 Fig. The standardized regression coefficients for the mediating effect of positive flexibility and the moderating effect of ERD on the relationship between discrimination and anxiety**. \**p* < .05, \*\**p* < .01, \*\*\**p* < .001.

**S3 Fig. The standardized regression coefficients for the mediating effect of negative flexibility and the moderating effect of ERD on the relationship between discrimination and anxiety**. \**p* < .05, \*\**p* < .01, \*\*\**p* < .001.

