## Supplementary figures and images for "Moderating the link between discrimination and adverse mental health outcomes: Examining the protective effects of cognitive flexibility and emotion regulation"

### Supplemental Figure 1

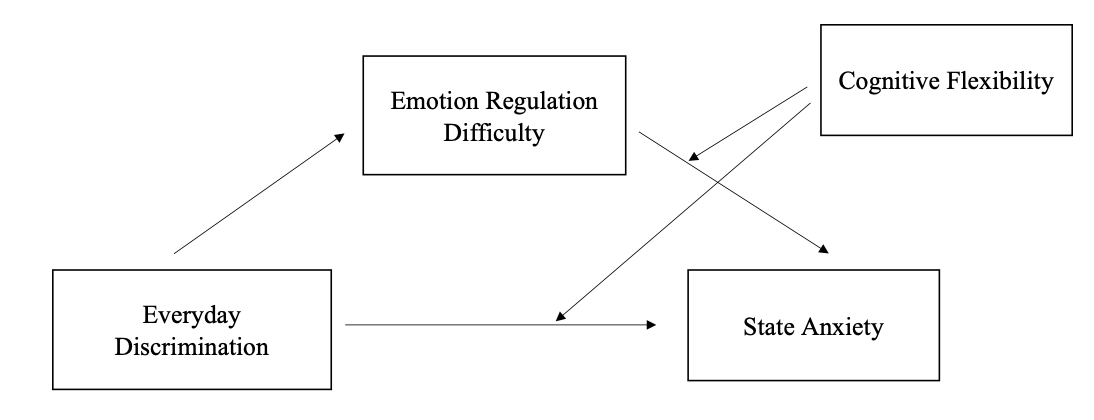

### Supplemental Figure 2

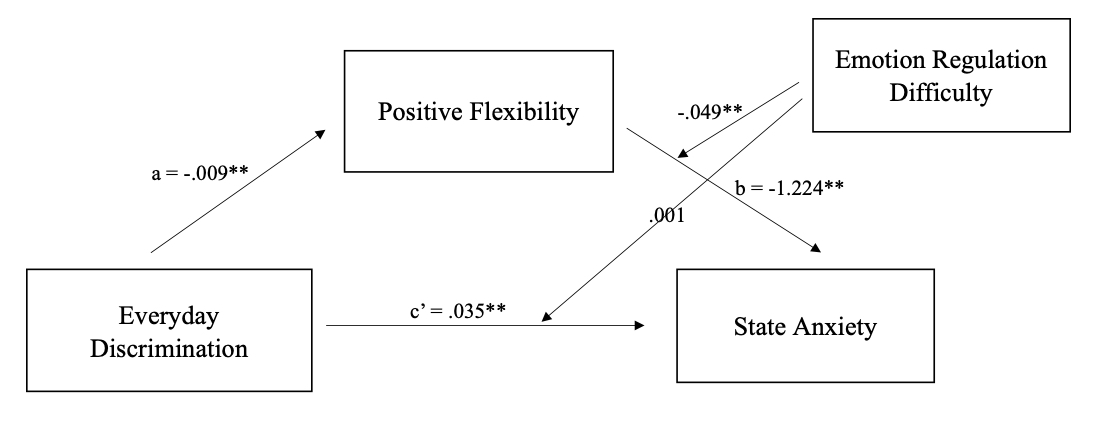

### Supplemental Figure 3

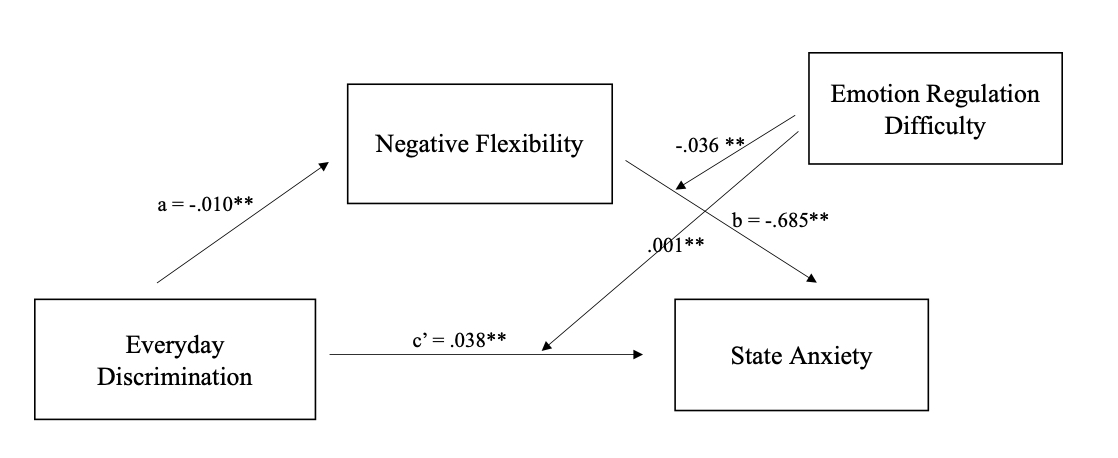
